# Development and validation of a machine learning model to predict cognitive behavioral therapy outcome in obsessive-compulsive disorder using clinical and neuroimaging data

**DOI:** 10.1101/2025.02.14.25322265

**Authors:** Laurens A. van de Mortel, Willem B. Bruin, Pino Alonso, Sara Bertolín, Jamie D. Feusner, Joyce Guo, Kristen Hagen, Bjarne Hansen, Anders Lillevik Thorsen, Ignacio Martínez-Zalacaín, Jose M Menchón, Erika L. Nurmi, Joseph O’Neill, John C. Piacentini, Eva Real, Cinto Segalàs, Carles Soriano-Mas, Sophia I. Thomopoulos, Dan J. Stein, Paul M. Thompson, Odile A. van den Heuvel, Guido A. van Wingen

## Abstract

Cognitive behavioral therapy (CBT) is a first-line treatment for obsessive-compulsive disorder (OCD), but clinical response is difficult to predict. In this study, we aimed to develop predictive models using clinical and neuroimaging data from the multicenter Enhancing Neuro-Imaging and Genetics through Meta-Analysis (ENIGMA)-OCD consortium.

Baseline clinical and resting-state functional magnetic imaging (rs-fMRI) data from 159 adult patients aged 18-60 years (88 female) with OCD who received CBT at four treatment/neuroimaging sites were included. Fractional amplitude of low frequency fluctuations, regional homogeneity and atlas-based functional connectivity were computed. Clinical CBT response and remission were predicted using support vector machine and random forest classifiers on clinical data only, rs-fMRI data only, and the combination of both clinical and rs-fMRI data.

The use of only clinical data yielded an area under the ROC curve (AUC) of 0.69 for predicting remission (p=0.001). Lower baseline symptom severity, younger age, an absence of cleaning obsessions, unmedicated status, and higher education had the highest model impact in predicting remission. The best predictive performance using only rs-fMRI was obtained with regional homogeneity for remission (AUC=0.59). Predicting response with rs-fMRI generally did not exceed chance level. Machine learning models based on clinical data may thus hold promise in predicting remission after CBT for OCD, but the predictive power of multicenter rs-fMRI data is limited.

## Introduction

Obsessive-compulsive disorder (OCD) is a psychiatric disorder with a lifetime prevalence of 2-3% [1] and is characterized by repetitive thoughts of an intrusive and distressing nature, and/or repetitive mental and behavioral compulsions. Current common treatment options for OCD involve cognitive behavioral therapy (CBT) with exposure and response prevention (ERP) or pharmacological treatment with a selective serotonin reuptake inhibitor [2, 3]. With ERP, individuals with OCD are exposed to their obsessions and subsequently taught to resist the urge of compulsive behavior and tolerate the associated distress. The aim is to diminish the associated emotional response, and the behaviors and avoidance done in attempts to reduce emotions, which thereby break the reinforcing cycle of obsessions and compulsive behaviors [4]. While approximately 50% of individuals with OCD benefit from ERP/CBT (hereafter referred to as CBT), they sometimes only achieve a partial reduction in symptoms, can result in dropout rates of 19%, and may not always be as cost effective as pharmacological treatment [2, 5–8]. It currently cannot be accurately predicted which patients will benefit from CBT and why. If treatment outcomes could be accurately predicted for individual patients, this could enable personalized treatment planning and improve our understanding of the factors underlying treatment response.

The use of machine learning may provide such opportunities. Predictive models can use both clinical and neuroimaging data on brain structure and function to identify (bio)markers relevant for predicting treatment outcomes. Meta-analyses have identified multiple clinical factors that are related to poorer CBT response at the group level, such as higher OCD symptom severity at baseline as measured by the Yale-Brown Obsessive Compulsive Scale (Y-BOCS), increased anxiety, higher age, comorbid personality disorder, and hoarding subtypes, but these factors cannot make accurate predictions for individual patients [9–13]. Machine learning studies have started to test multivariate predictive models based on clinical factors, but the accuracy of those models has been limited [14, 15]. In an attempt to improve model accuracy and uncover biomarkers of CBT response, machine learning studies have incorporated functional magnetic resonance imaging (fMRI) data. Initial studies indeed suggest that predictive models using fMRI data are more accurate than models based on clinical data [16–18]. However, those studies are limited by the use of smaller samples (N<60) from single research sites, which tend to yield inflated model accuracy and decreased generalizability to other samples, due to overfitting to features of the data they are trained on [19, 20]. To obtain more robust biomarkers, large multicenter data are required with independent validation methods. Currently, it is unclear whether CBT outcome can be predicted in multicenter datasets and whether clinical data, fMRI data, or its combination yields the highest accuracy for predicting clinical outcome.

In this study, we predicted CBT outcomes in OCD using pre-treatment 1) clinical and demographic data, and 2) resting-state fMRI data to estimate brain function using derivatives that have been associated with OCD pathophysiology (i.e. fALFF, ReHo, and functional connectivity [21–23]. Data were obtained from several sites of the multicenter Enhancing Neuro-Imaging and Genetics through Meta-Analysis (ENIGMA) OCD consortium. We trained machine learning models to predict clinical response, remission, and post-treatment symptom severity as determined by the Y-BOCS, and evaluated model accuracy in independent samples using leave-one-site-out cross-validation. The study is reported in accordance with TRIPOD guidelines for diagnostic studies [24].

## Methods

### Participants

The initial sample consisted of 300 participants for whom rs-fMRI data and information about CBT outcome was available. We excluded participants below 18 years of age (n=56), samples from sites with N<20 [25] to ensure classifiers were provided with sufficient data per site (n=71), and 14 participants with insufficient data quality (rotation/translation>4 mm/degrees, average FD>0.25 with <100 volumes), leading to a sample of 159 participants (88 female, mean age 33±9.5 years) across four ENIGMA-OCD neuroimaging sites [18, 26, 27]. OCD was diagnosed according to the diagnostic criteria from the Diagnostic and Statistical Manual for Mental Disorders IV or 5 (DSM-IV/5). All studies were approved by the local institutional review board and participants provided written informed consent.

Although all sites administered CBT focused on ERP, exact CBT protocols differed across sites. One site administered the Bergen 4-day treatment protocol. The three other sites administered CBT through standard protocols, with a varying number of sessions and duration. Two of these three sites administered CBT in a group setting. All sites included homework tasks as an additional part of the therapy.

### Clinical data

At baseline, clinical and demographic data (henceforth termed clinical data) were recorded and consisted of the participants’ age, biological sex, education level, medication use, current diagnosis of depression, current diagnosis of an anxiety disorder, Y-BOCS at baseline, and obsession type (aggressive, cleaning/contamination, sexual/religious, hoarding, and/or ordering/symmetry obsessions). For an overview of all clinical data, see Table 1.

**Table 1.** Demographic and clinical data of the total participant sample (N=159).

| Variable | Mean±SD/N |
| --- | --- |
| Age | 33.0±9.5 |
| Sex | 88 female/71 male |
| Education (yrs) | 13.8±3.0 |
| Medicated | 101 prior/129 during |
| Current diagnosis of major depressive disorder | 29 |
| Current diagnosis of an anxiety disorder | 42 |
| Y-BOCS | 26.3±4.8 |
| Aggression/checking obsessions | 134 |
| Cleaning/contamination obsessions | 99 |
| Sexual/religious obsessions | 70 |
| Hoarding obsessions | 52 |
| Ordering/symmetry obsessions | 52 |
| Clinical response ($\geq 35\%$ reduction) | 110 |
| Remission (Y-BOCS$\leq 12$) | 67 |

### Neuroimaging data

Resting-state fMRI (rs-fMRI) scans were acquired (see Table S1 for imaging acquisition parameters) and processed locally using the fMRIPrep-based Harmonized AnaLysis of Functional MRI pipeline (HALFpipe) [28–30], according to standardized protocols (see http://enigma.ini.usc.edu/protocols/functional-protocols/) as described in Bruin et al., 2023. Preprocessing steps included motion correction, slice timing and susceptibility distortion correction (if available), normalization, and denoising using grand mean scaling with a mean value of 10,000, and correction of head motion, white matter, and cerebrospinal fluid artifacts using the top five principal noise components in aCompCor and ICA-AROMA [31].

To estimate local brain activity, fMRI data were band-pass filtered (0.01-0.1 Hz) and fALFF and ReHo were extracted, which measure the local spontaneous neural activity and its regional coherence, respectively [32]. These values were subsequently smoothed with a 6-mm FWHM kernel. Voxel-wise values were subsequently averaged per region of interest (ROI) to obtain 400 mean fALFF and ReHo values based on the Schaefer 400 atlas [33].

For brain-wide functional connectivity, fMRI data were high-pass filtered (0.008 Hz). Since ROI time series with less than 80% voxel coverage were excluded during data extraction, we restricted the sample for the connectivity analysis by excluding subjects with >20% missing ROIs (n=39). The remaining correlation matrices (n=120) were then masked to include only regions that had coverage for all subjects, leading to a 330-by-330 connectivity matrix with regions from the Schaefer 400-17 network atlas [33], 17 ROIs from the subcortical Harvard-Oxford Atlas [34], and 17 cerebellar ROIs from the Buckner 17-network atlas [35].

### Machine learning

For binary classification, we predicted two types of CBT outcome for each data modality: clinical response (defined as ≥35% reduction in Y-BOCS) and remission (≤Y-BOCS of 12) [36]. Additionally, we performed regression on post-treatment Y-BOCS to overcome limitations of dichotomizing continuous Y-BOCS using support vector regression and RF regressor with identical parameters on the grid search as for binary classification.

Training and validation were performed with a nested loop, in which the model was trained on three sites and validated on the fourth independent site. We compared the performance of random forest (RF) and support vector machine (SVM) models on predicting CBT outcome with clinical data only, rs-fMRI data only, or different combinations of clinical and rs-fMRI data. For each of the four folds, label-stratified grid search was performed on the training data to find the optimal hyperparameters for SVM (C: 0.1-1000, gamma: 0.0001-1, kernel: radial basis function or linear) and RF (maximum number of features: 10-300, minimum samples per leaf: 1-10, minimum samples per node split: 2-20, number of decision trees: 100-1000) with balanced accuracy as the scoring function. These hyperparameters were subsequently used in the model to predict outcome in the held out test site. If there was class imbalance for a CBT outcome variable (>60% belonging to the majority class), random under-sampling of the majority class was performed on the training data.

We also performed an additional classification using nested 3×5 cross-validation with five site-stratified outer folds and three CBT-outcome stratified inner folds. Because multi-site imaging data has been shown to induce noise and biases that counteract the learning of relevant features in shuffled cross-validations [37], we scaled and fitted the data on the training and testing set separately and performed ComBat [38] regression to regress out batch/scanner effects of the different imaging sites on the train and test set separately for the outer folds.

Model performance was assessed by averaging the area under the receiver operating characteristic curve (AUC), positive predictive value (PPV), negative predictive value (NPV), sensitivity, and specificity over the different sites/folds for classification. We obtained 95% confidence intervals for AUC values using an analytical computation of the DeLong method [39]. Root mean square error (RMSE) and coefficient of determination (R2) over the different sites/folds were calculated for regression. Statistical significance of the best performing model was statistically tested with 1000 permutations, and Shapley Additive explanation (SHAP) values were extracted for model interpretation [40].

### Univariate analysis

Besides the multivariate analyses, we performed confirmatory univariate analyses for both the clinical and rs-fMRI data. A whole-brain univariate analysis was performed to compare differences in fALFF and ReHo data between remitters and non-remitters while correcting for covariates of age, biological sex, medication use, and imaging site with a two-sample *t*-test using Statistical Parametric Mapping 12 (SPM12, https://www.fil.ion.ucl.ac.uk/) in Matlab R2018b [41]. Multiple comparisons correction of whole brain voxel-wise comparisons was employed with family-wise-error (FWE) rate correction at α=0.05 on the cluster level (cluster forming threshold p<0.001). Connectivity matrices were compared between remitters and non-remitters with the Network Based Statistics (NBS) toolbox in Matlab R2018b using 5000 permutations at α=0.05 (network based statistics method, significance based on cluster intensity) while correcting for age, biological sex, medication use, and imaging site.

## Results

### Patient characteristics

Participants had a mean Y-BOCS of 26.3±4.8 at baseline, indicating severe OCD. On average, participants received 16.0±6.6 sessions of CBT with an average treatment duration of 11.5±8.9 weeks. Following treatment, Y-BOCS significantly reduced to 14.7±6.7; t(158)=22.16,p<0.001). The majority of the 159 individuals (110, 69%) responded to the treatment (≥35% reduction in Y-BOCS) and 67 (42%) achieved remission (Y-BOCS ≤12). Patient characteristics are described in Table 1.

### Classification Performance

#### Clinical data only

Performance metrics across all data modalities and outcome predictions are depicted in Figure 1,2, S1 and S2.

**Figure 1:**
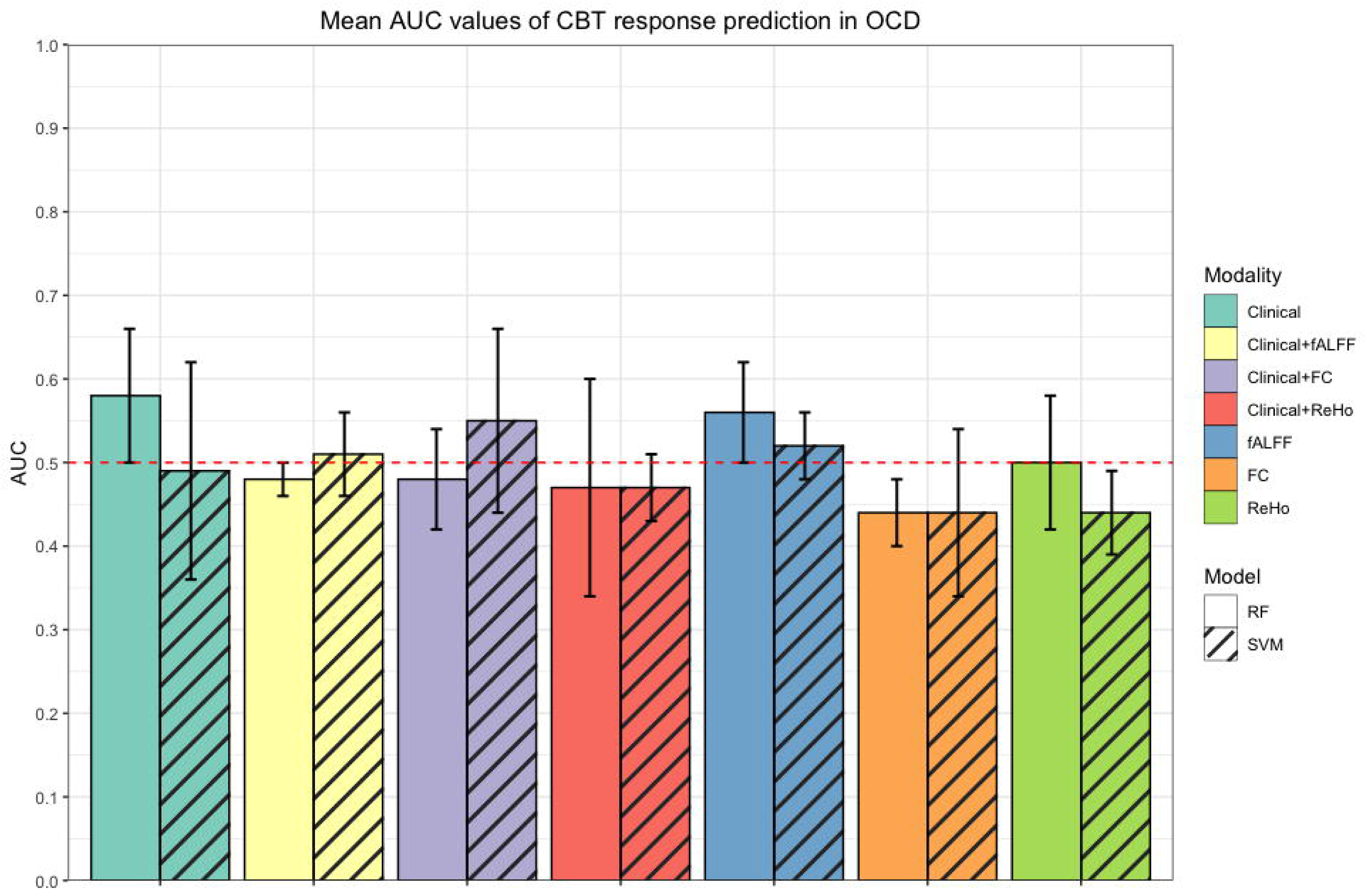
Mean AUC values of SVM and RF in the prediction of CBT response (≥35% reduction in Y-BOCS) in OCD. Mean and standard deviations depicted for each AUC, modality, and model. Clinical=clinical data, fALFF=fractional amplitude of low frequency fluctuations, ReHo=regional homogeneity, FC=functional connectivity, RF=random forest, SVM=support vector machine.

**Figure 2:**
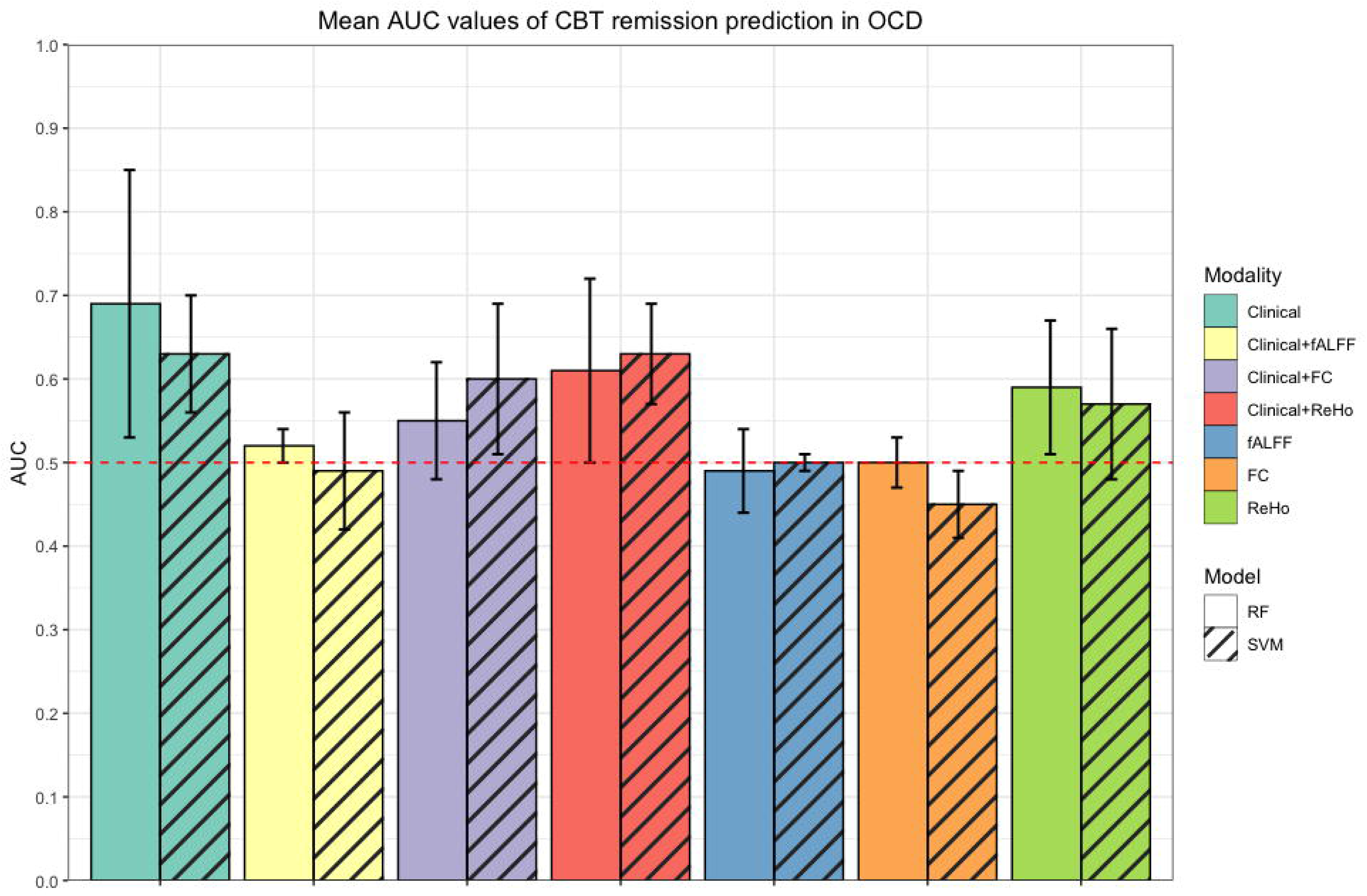
AUC values of SVM and RF in the prediction of CBT remission (Y-BOCS≤12) in OCD. Mean and standard deviations depicted for each AUC, modality, and model. Clinical=clinical data, fALFF=fractional amplitude of low frequency fluctuations, ReHo=regional homogeneity, FC=functional connectivity, RF=random forest, SVM=support vector machine.

Multivariate prediction of response after CBT using clinical data yielded a low mean AUC of 0.58 (see Tables 2 and 3). The prediction of remission achieved the highest performance with a mean AUC of 0.69 using a random forest classifier (95% CI [0.58, 0.73], p=0.001). From this model, the variables with the highest SHAP values indicated that a lower Y-BOCS at baseline, lower age, an absence of cleaning obsessions and unmedicated status, and higher educational level contributed most to a prediction of remission (see Figure 3).

**Figure 3:**
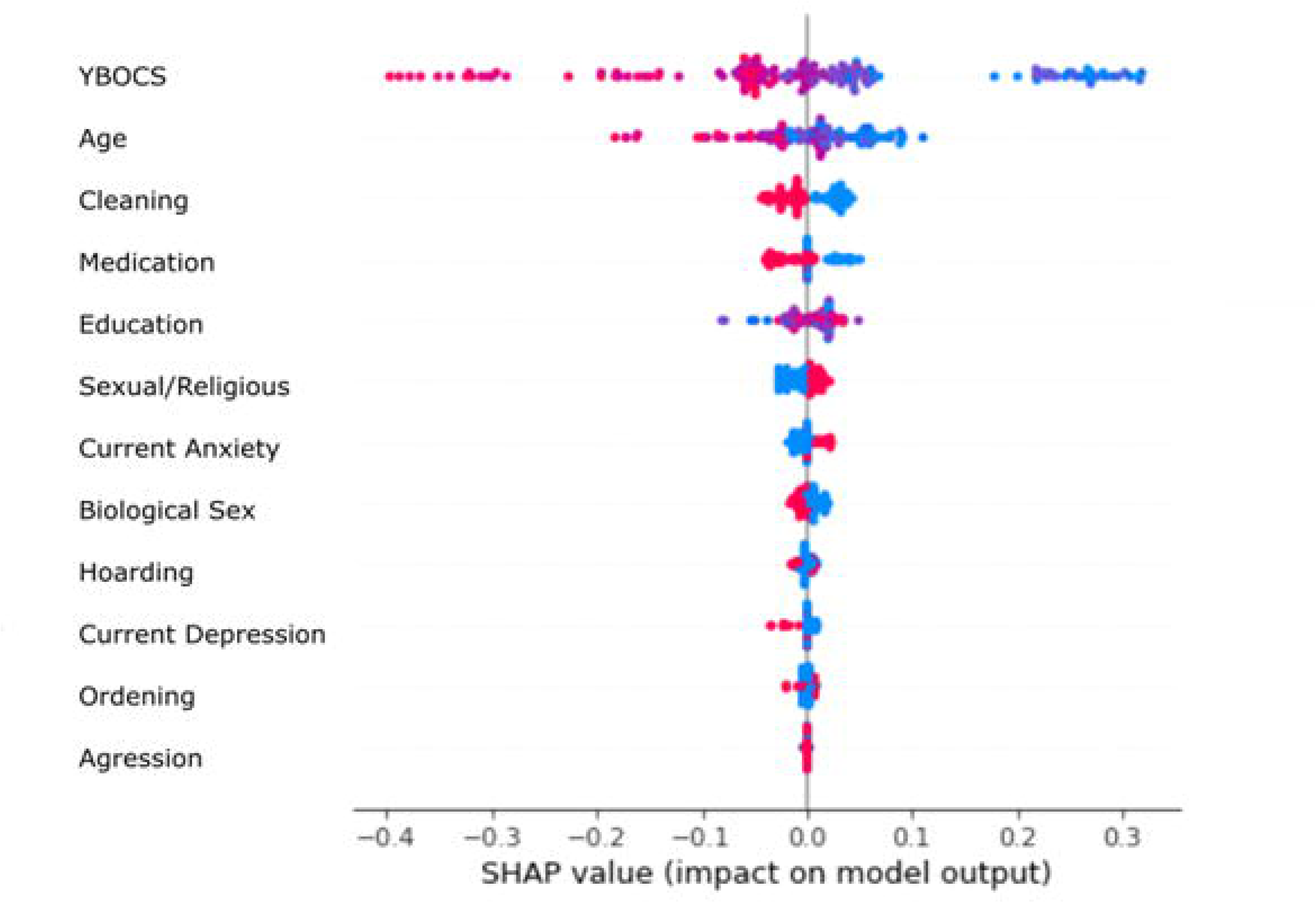
SHAP values of all clinical features in predicting CBT remission, indicating that lower Y-BOCS severity, lower age, an absence of cleaning obsessions, unmedicated status, and higher education are the most relevant features for predicting remission.

**Table 2:** SVM and RF prediction performance of CBT response (. ≥**35% reduction) in OCD with rs-fMRI and clinical characteristics in a leave-one-site-out framework.**

|  | fALFF | ReHo | Functional<br>Connectivity | Clinical<br>data | Clinical<br>+fALFF | Clinical<br>+ReHo | Clinical<br>+Functional<br>Connectivity |
| --- | --- | --- | --- | --- | --- | --- | --- |
| **SVM** |  |  |  |  |  |  |  |
| AUC (95%<br>CI) | 0.52±0.04<br>(0.41-0.58) | 0.44±0.05<br>(0.35 –<br>0.52) | 0.44±0.10<br>(0.31 – 0.50) | 0.49±0.13<br>(0.43 –<br>0.60) | 0.51±0.05<br>(0.43 –<br>0.60) | 0.47±0.04<br>(0.37 –<br>0.53) | 0.55±0.11<br>(0.44 – 0.62) |
| PPV | 0.33±0.33 | 0.51±0.17 | 0.54±0.36 | 0.69±0.05 | 0.53±0.32 | 0.45±0.27 | 0.43±0.11 |
| NPV | 0.37±0.15 | 0.29±0.09 | 0.25±0.16 | 0.41±0.29 | 0.32±0.19 | 0.29±0.10 | 0.66±0.14 |
| Sensitivity | 0.44±0.44 | 0.33±0.27 | 0.34±0.25 | 0.60±0.22 | 0.54±0.34 | 0.34±0.34 | 0.57±0.15 |
| Specificity | 0.61±0.41 | 0.56±0.19 | 0.54±0.37 | 0.39±0.28 | 0.48±0.34 | 0.59±0.30 | 0.53±0.11 |
| **RF** |  |  |  |  |  |  |  |
| AUC (95%<br>CI) | 0.56±0.06<br>(0.45 –<br>0.62) | 0.50±0.08<br>(0.47 –<br>0.64) | 0.44±0.04<br>(0.35 – 0.54) | 0.58±0.08<br>(0.50 –<br>0.66) | 0.48±0.02<br>(0.44 –<br>0.61) | 0.47±0.13<br>(0.45 –<br>0.62) | 0.48±0.06<br>(0.38 – 0.56) |
| PPV | 0.73±0.10 | 0.68±0.12 | 0.56±0.10 | 0.75±0.04 | 0.65±0.12 | 0.65±0.13 | 0.36±0.09 |
| NPV | 0.36±0.16 | 0.34±0.15 | 0.29±0.09 | 0.45±0.23 | 0.32±0.11 | 0.31±0.18 | 0.59±0.09 |
| Sensitivity | 0.65±0.12 | 0.52±0.19 | 0.35±0.20 | 0.66±0.16 | 0.40±0.24 | 0.49±0.17 | 0.43±0.08 |
| Specificity | 0.47±0.22 | 0.49±0.21 | 0.54±0.18 | 0.50±0.16 | 0.57±0.25 | 0.44±0.26 | 0.53±0.11 |

**Table 3:** SVM and RF prediction performance of CBT remission (Y-BOCS ≤12) in OCD with rs-fMRI and clinical data in a leave-one-site-out framework.

|  | fALFF | ReHo | Functional Connectivity | Clinical data | Clinical +fALFF | Clinical +ReHo | Clinical +Functional Connectivity |
| --- | --- | --- | --- | --- | --- | --- | --- |
| **SVM** |  |  |  |  |  |  |  |
| AUC (95% CI) | 0.50±0.01<br>(0.48-0.64) | 0.57±0.09<br>(0.53-0.68) | 0.45±0.04<br>(0.37 – 0.56) | 0.63±0.07<br>(0.56 – 0.71) | 0.49±0.07<br>(0.42 – 0.58) | 0.63±0.06<br>(0.55 – 0.70) | 0.60±0.09<br>(0.48 – 0.65) |
| PPV | 0.33±0.21 | 0.44±0.28 | 0.31±0.11 | 0.55±0.06 | 0.34±0.20 | 0.53±0.10 | 0.50±0.13 |
| NPV | 0.48±0.28 | 0.65±0.02 | 0.44±0.26 | 0.72±0.18 | 0.63±0.19 | 0.77±0.14 | 0.69±0.13 |
| Sensitivity | 0.40±0.38 | 0.34±0.24 | 0.47±0.34 | 0.71±0.13 | 0.30±0.38 | 0.71±0.23 | 0.42±0.28 |
| Specificity | 0.60±0.37 | 0.81±0.07 | 0.42±0.29 | 0.55±0.18 | 0.67±0.28 | 0.55±0.12 | 0.77±0.13 |
| **RF** |  |  |  |  |  |  |  |
| AUC (95% CI) | 0.49±0.05<br>(0.40 – 0.56) | 0.59±0.08<br>(0.50-0.64) | 0.50±0.03<br>(0.44 – 0.55) | 0.69±0.16<br>(0.58 – 0.73) | 0.52±0.02<br>(0.43 – 0.59) | 0.61±0.11<br>(0.50 – 0.64) | 0.55±0.07<br>(0.49 – 0.66) |
| PPV | 0.43±0.36 | 0.55±0.13 | 0.21±0.22 | 0.67±0.20 | 0.51±0.37 | 0.56±0.10 | 0.39±0.25 |
| NPV | 0.60±0.10 | 0.65±0.11 | 0.62±0.07 | 0.75±0.20 | 0.62±0.11 | 0.66±0.16 | 0.66±0.07 |
| Sensitivity | 0.27±0.38 | 0.40±0.17 | 0.09±0.09 | 0.70±0.21 | 0.33±0.38 | 0.42±0.25 | 0.27±0.23 |
| Specificity | 0.72±0.35 | 0.79±0.02 | 0.91±0.06 | 0.68±0.26 | 0.72±0.38 | 0.79±0.05 | 0.83±0.09 |

#### Neuroimaging data only

Mean AUCs for predicting clinical response and remission using fALFF, ReHo, and functional connectivity data ranged between 0.44 to 0.59 (see Table 2 and 3 for all performances across the rs-fMRI measures).

ReHo achieved the highest performance in predicting remission, with a mean AUC of 0.59 using an SVM, although prediction of clinical response using ReHo only achieved a mean AUC of 0.50. fALFF predicted response and remission with mean AUC values ranging between 0.49 to 0.56 for predicting remission and response with an RF, respectively. Functional connectivity had the lowest performance of all imaging measures with mean AUC values ranging between 0.44 to 0.50 for predicting response and remission with RF.

#### Multimodal data

We next evaluated whether the combination of clinical and rs-fMRI data could result in better predictions. The use of multimodal data did not outperform the use of single rs-fMRI measures or clinical data in the prediction of CBT outcome: the best performing model was the combination of clinical data and ReHo with a mean AUC of 0.63 for predicting remission using an SVM.

#### 5-fold cross validation

To evaluate whether models could perform better when data from every site is available during training, we additionally performed 5-fold cross-validation with participants across all sites shuffled over the folds (see Table S2 and S3). Compared to leave-one-site-out cross validation, 5-fold cross-validation yielded similar or marginally higher prediction performances for functional connectivity (AUC=0.55) and the combination of clinical data and fALFF (AUC=0.59), but none of the modalities and models outperformed the best model with leave-one-site-out cross-validation.

#### Regression performance

We next evaluated whether post-treatment Y-BOCS could be accurately predicted using SVR and RF Regressor. This generally yielded poor results with high RMSE and low R2 values, especially for the rs-fMRI data. The use of clinical data for predicting post-treatment Y-BOCS using an RF had the relatively lowest mean RMSE of 6.05 (see Table S4).

#### Univariate analysis

Finally, we evaluated whether there were univariate associations between baseline differences in ReHo, fALFF, functional connectivity and clinical data between remitters and non-remitters. We found no statistically significant differences between both groups in any of the imaging measures.

Clinically, remitters only showed lower baseline Y-BOCS severity (M=24.2,sd=4.5) than non-remitters (M=27.8, sd=4.5), t(142)=4.9, p=<0.001, Bonferroni corrected). To explore whether baseline Y-BOCS could also predict remission, we computed the ROC curve for the entire sample. This yielded a total AUC of 0.72 for predicting remission. This analysis also revealed a Y-BOCS cut-off point of 23.5 with the best balanced accuracy in predicting remission (balanced accuracy: 0.67), albeit with a poor balance between sensitivity (0.87) and specificity (0.46). For all Y-BOCS cut-off points and their respective performance in predicting remission, see Table S5.

## Discussion

In this multicenter ENIGMA-OCD cohort study, we investigated the potential of using baseline clinical data and fALFF, ReHo, and functional connectivity measures from resting-state fMRI data for predicting CBT outcomes in adult participants diagnosed with OCD. We found moderately positive results in the prediction of CBT remission using clinical data (AUC=0.69), but also found that prediction of CBT outcome with only rs-fMRI data was unsuccessful: mean AUC values for various rs-fMRI features ranged between 0.44 (for predicting CBT response with functional connectivity data) to 0.59 (for predicting CBT remission with ReHo data). In general, performance was better for predicting CBT remission than response, even when using random under-sampling to account for the class imbalance of clinical response. We also attempted regression on the post-treatment Y-BOCS value, which also yielded unsatisfactory performance for both rs-fMRI and clinical data (high RMSE and low R2 values).

We achieved the highest performance with clinical data, but only for the prediction of remission. With an AUC of 0.69 with leave-one-site-out cross validation, we reach a performance that falls just short of being classified as acceptable discrimination [42], but this performance is higher than reported in previous work on predicting CBT outcome in OCD with clinical data [14, 15]. The performance may have been limited due to inter-site differences in CBT protocols and patient inclusion. One of the four sites followed the Bergen 4 day CBT protocol, which has shown high efficacy in the treatment of OCD [43] regardless of pre-treatment Y-BOCS severity [44]. The other sites followed different ERP protocols in both group and individual settings, with varying number of sessions, duration, and efficacy. Despite these differences, our results show that the factors determining CBT remission are relatively universal: lower baseline Y-BOCS, lower age, an absence of cleaning obsessions, unmedicated status, and a higher education increase the chances of being classified as a remitter. Feature importance in machine learning models should be interpreted with caution as the models have a multivariate nature, but previous studies have consistently indicated that high Y-BOCS at baseline predicts worse CBT outcome, which is corroborated by our study [9, 45]. Our ROC analysis revealed that remission indeed could also be predicted with the baseline Y-BOCS alone with a comparable mean performance to that of our multivariate models. However, while this yielded a high sensitivity (0.87) for the highest balanced accuracy (0.67) with a cut-off Y-BOCS point of 23.5, specificity was only 0.46. This indicates that Y-BOCS alone cannot predict non-remission, and that a better balance between sensitivity and specificity can be achieved by using multivariate models and additional clinical variables, besides baseline Y-BOCS.

The importance of age and educational level for CBT outcome have also been reported previously, although not consistently [11, 46]. Contrary to prior studies, we found no evidence for the hoarding obsession subtype being negatively associated with CBT outcome [9, 10]. Instead, there was an indication that patients with contamination obsessions were less likely to remit. While studies have shown that contamination obsessions can be treated successfully with CBT [47, 48], these studies tend to focus on clinical response, which may not necessarily extend to clinical remission for this subtype.

In general, the prediction of CBT response did not exceed chance-level when rs-fMRI and clinical data were used jointly. The fact that response could not be predicted successfully for pooled rs-fMRI and clinical data may lie in the underlying data distributions of CBT outcomes. As most of the participants achieved a clinical response to CBT, there was a large class imbalance between groups, which despite undersampling of the majority class made prediction difficult. For remission, we predicted an outcome that was more balanced and may stand out more in a sample where the majority achieved response, but a minority achieved remission. The use of a clinical decision model that predicts remission may also be more beneficial as patients achieving remission are less likely to relapse [49], but whether a model performance of 0.69 AUC is actually beneficial to patient care will need to be investigated in a thorough cost-benefit analysis [50].

The unsuccessful prediction of CBT outcome with rs-fMRI is not in line with earlier research which has reported that, at least in smaller monocenter samples, rs-fMRI may have a potential in predicting CBT outcome through baseline activity and connectivity of subcortical and cortical areas such as the ventromedial prefrontal cortex and subcortex [16–18]. While these studies show that functional connectivity may be relevant for the prediction of CBT response for individual institutes, the chance level performance in our study indicates that such models cannot generalize to data from other institutes.

The use of multi-site data provides opportunities to increase the sample size and thereby the generalizability of model performance, but this variation and heterogeneity could also negatively impact model performance. Increases in sample sizes in psychiatric research tend to increase data heterogeneity and thereby reduce model performance [20, 51], which proves even more difficult when considering that OCD has a highly heterogeneous biological and clinical presentation [52–54], and large samples are often obtained by the use of multiple scanners at different imaging sites which additionally induces artificial variability [55, 56]. Further variability in the biological data in this study includes the use of medication, which has been shown to affect fMRI signals [57–59]. In our 5-fold cross validation analysis, we employed ComBat on the rs-fMRI data to mitigate between-site variability. However, this unfortunately did not improve performance as compared to leave-one site out cross-validation, which could imply that no observable biological markers of therapy response were present in the baseline data. This notion is supported by our univariate statistical analyses where we found no baseline differences in fALFF, ReHo, and functional connectivity between future remitters and non-remitters. While multivariate machine learning analyses are typically more sensitive to detect patterns in neuroimaging data than univariate analyses [60], the results from this univariate analysis indicate a possibility that no useful biological markers of brain activity related to CBT outcome were present, at least among those selected, in the rs-fMRI data for our models.

In light of these findings, the strengths and limitations of this study should be considered. The relatively large multi-site sample size for both neuroimaging and clinical data is a strength of our study, allowing for better representation of the large clinical heterogeneity in OCD and improvement of model generalizability. Although we almost reach acceptable discrimination for predicting remission with the use of clinical data only, the sample size in this study may still have been too limited for reliable model performance [61], especially with the use of neuroimaging. Unfortunately, larger sample sizes also increase the number of confounding factors that are difficult to account for, and this is a limitation of our study: there were site differences in both the rs-fMRI acquisition and CBT protocols with variations in treatment type, duration, and efficacy. Many of the patients were also simultaneously taking psychotropic medication. Without accounting for these factors, no definitive conclusion can yet be drawn about the value of rs-fMRI data for the prediction of CBT outcome in OCD.

In summary, this study used multi-site imaging and clinical data from a relatively large (n=159) sample of individuals from the ENIGMA-OCD cohort in an attempt to find reliable biomarkers of CBT response in OCD. We showed moderate performance in the prediction of remission with the use of clinical data. Baseline YBOCS severity, age, education level, unmedicated status and an absence of cleaning obsessions were the most relevant features to achieve remission. The potential for clinical use needs to be further evaluated before these results can be implemented. Yet, our study did not reveal any useful biomarkers of CBT outcome derived from resting-state fMRI data. While this study has limitations that prevent us from drawing any definite conclusions on the use of rs-fMRI data in predicting CBT outcome, our results imply that clinical data are more relevant for the prediction of CBT remission in OCD.

## Data Availability

Code and data used in this work are available upon reasonable request.

## Acknowledgements

For full information regarding grant support to the ENIGMA-OCD Working Group, see https://enigma.ini.usc.edu/about-2/funding/.

This study specifically is supported by grants awarded by the National Institute of Mental Health (NIMH; Grant No. R01MH081864; J.O. and J.C.P., R01MH085900; J.O. and J.F, R01MH129742; P.T., K23MH094613; E.N.), the National Institute on Aging Research Project (Grant No. R01AG058854; P.T.), The Western Norway Regional Health Authority (Grant. No. 911754 and 911880; A.L.T.), the South African Medical Research Council (D.S.), the Carlos III Health Institute ( Grant No. PI19/01171;C.S.M, Grant No. CM21/00278 Co Funded by the European Social fund; S.B.), the Department of Health, Generality of Catalonia (PERIS SLT006/17/249; C.S.M.); The Agency for Management of University and Research Grants (2021-SGR-01017; C.S.M.), The María de Maeztu Unit of Excellence, Institute of Neurosciences, University of Barcelona (CEX2021-001159-M; C.S.M) and the International OCD Foundation (Innovator Award; O.V.D.H).

