## Supplementary Information for "Development and validation of a machine learning model to predict cognitive behavioral therapy outcome in obsessive-compulsive disorder using clinical and neuroimaging data"

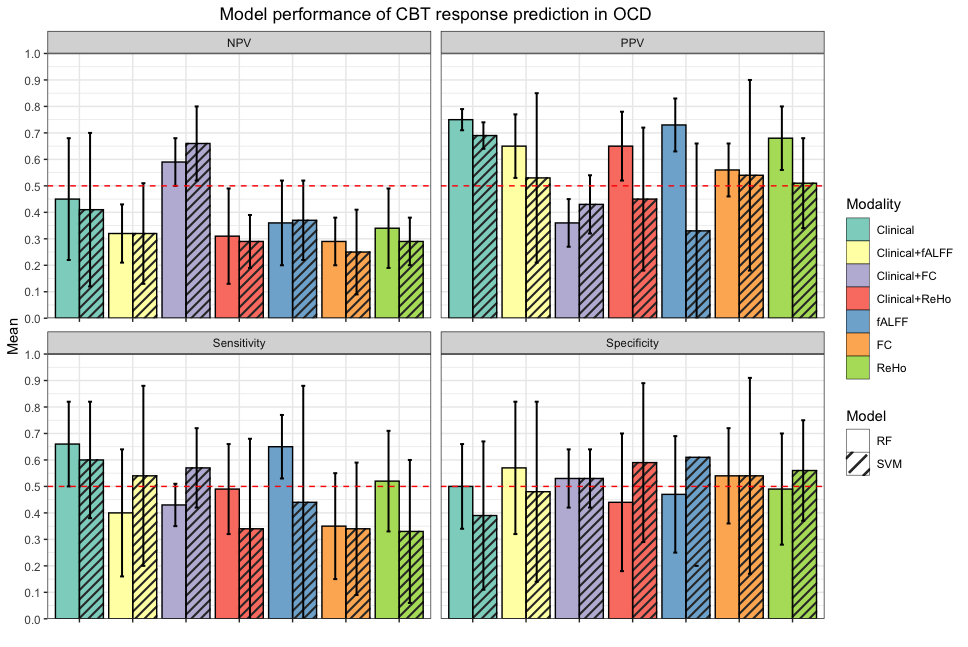

Figure S1: Performance metrics of SVM and RF in the prediction of CBT response (≥35% reduction in Y-BOCS) in OCD. Mean and standard deviations depicted for each metric, modality, and model. Clinical=clinical data, fALFF=fractional amplitude of low frequency fluctuations, ReHo=regional homogeneity, FC=functional connectivity, RF=random forest, SVM=support vector machine, NPV=negative prediction value, PPV=positive prediction value.

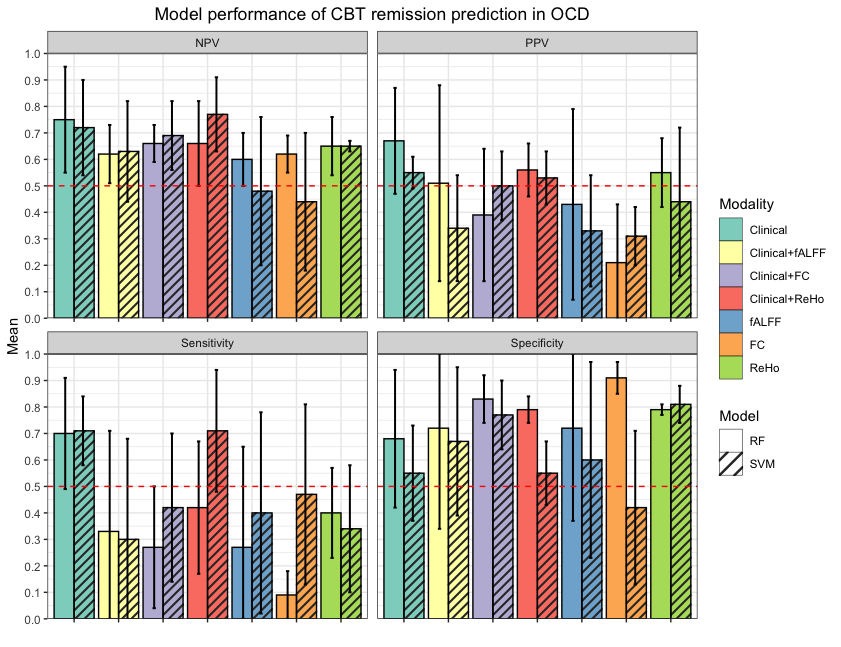

Figure S2: Performance metrics of SVM and RF in the prediction of CBT remission (Y-BOCS≤12) in OCD. Mean and standard deviations depicted for each metric, modality, and model. Clinical=clinical data, fALFF=fractional amplitude of low frequency fluctuations, ReHo=regional homogeneity, FC=functional connectivity, RF=random forest, SVM=support vector machine, NPV=negative prediction value, PPV=positive prediction value.

Table S1: Imaging acquisition parameters per imaging site.

| Site | Barcelona (ANTIGA study) | Barcelona (COMPULSE study) | Bergen | UCLA |
| --- | --- | --- | --- | --- |
| Scanner | GE Signa Excite 1.5T | Philips Ingenia 3T | GE Discovery MR750 3T | Siemens Trio 3T |
| Voxel Size (mm) | 3.75x3.75x5 | 3x3x3 | 3.4x3.4x3.3 | 3x3x3 |
| TR (ms) | 2000 | 2000 | 1800 | 2000 |
| TE (ms) | 50 | 25 | 30 | 25 |
| Flip Angle | 90 | 90 | 80 | 78 |
| No. of volumes | 120 | 240 | 160 | 208 |

Table S2: SVM and RF performance of predicting CBT response (≥35% reduction) using rs-fMRI and clinical data with 5-fold cross-validation

|  | fALFF | ReHo | Functional Connectivity | Clinical data | Clinical+fALFF | Clinical+ReHo | Clinical+Functional Connectivity |
| --- | --- | --- | --- | --- | --- | --- | --- |
| **SVM** |  | | | | | | |
| AUC | 0.46±0.03 (0.41 – 0.57) | 0.53±0.06 (0.44 – 0.61) | 0.49±0.02  (0.44 – 0.63) | 0.48±0.05 (0.39 – 0.56) | 0.54±0.04 (0.47 – 0.63) | 0.48±0.04 (0.44 – 0.55) | 0.43±0.05 (0.31 – 0.49) |
| PPV | 0.64±0.10 | 0.57±0.30 | 0.55±.0.29 | 0.39±0.33 | 0.44±0.36 | 0.68±0.09 | 0.49±0.25 |
| NPV | 0.23±0.12 | 0.27±0.19 | 0.23±0.21 | 0.20±0.18 | 0.36±0.08 | 0.24±0.16 | 0.14±0.12 |
| Sensitivity | 0.57±0.31 | 0.66±0.35 | 0.64±0.35 | 0.45±0.40 | 0.44±0.34 | 360.85±0.10 | 0.59±0.30 |
| Specificity | 0.35±0.30 | 0.39±0.36 | 0.34±0.37 | 0.50±0.45 | 0.64±0.31 | 0.11±0.06 | 0.28±0.38 |
| **RF** |  | | | | | | |
| AUC | 0.46±0.02 (0.43 – 0.49) | 0.51±0.05 (0.47 – 0.55) | 0.50±0.07 (0.45 – 0.55) | 0.51±0.15 (0.43 – 0.58) | 0.49±0.06 (0.45 – 0.54) | 0.49±0.03 (0.45 – 0.52) | 0.54±0.09 (0.46 – 0.56) |
| PPV | 0.68±0.04 | 0.70±0.08 | 0.68±.0.06 | 0.69±0.11 | 0.69±0.08 | 0.69±0.06 | 0.69±0.13 |
| NPV | 0.05±0.10 | 0.45±0.46 | 0.40±0.49 | 0.32±0.09 | 0.23±0.29 | 0.27±0.39 | 0.18±0.26 |
| Sensitivity | 0.90±0.03 | 0.96±0.06 | 0.93±0.06 | 0.74±0.08 | 0.93±0.08 | 0.94±0.04 | 0.94±0.06 |
| Specificity | 0.03±0.05 | 0.07±0.06 | 0.06±0.08 | 0.29±0.09 | 0.05±0.07 | 0.03±0.04 | 0.13±0.19 |

Table S3: 5-fold SVM prediction performance of CBT remission (Y-BOCS ≤12) in OCD with rs-fMRI and clinical data with 5-fold cross-validation.

|  | fALFF | ReHo | Functional Connectivity | Clinical data | Clinical+fALFF | Clinical +ReHo | Clinical +Functional Connectivity |
| --- | --- | --- | --- | --- | --- | --- | --- |
| **SVM** |  | | | | | | |
| AUC | 0.47±0.08 (0.38 – 0.54) | 0.57±.0.09 (0.48 – 0.64) | 0.53±0.06 (0.46 – 0.64) | 0.64±0.05 (0.56 – 0.72) | 0.50±0.08 (0.41 – 0.57) | 0.59±0.06 (0.51 – 0.66) | 0.60±0.07 (0.48 – 0.66) |
| PPV | 0.40±0.14 | 0.49±0.16 | 0.42±0.08 | 0.57±0.07 | 0.41±0.12 | 0.50±0.07 | 0.49±.0.07 |
| NPV | 0.53±0.07 | 0.66±0.12 | 0.53±0.28 | 0.72±0.06 | 0.60±0.11 | 0.70±0.17 | 0.68±0.14 |
| Sensitivity | 0.56±0.10 | 0.57±0.21 | 0.57±0.19 | 0.64±0.15 | 0.48±0.26 | 0.59±0.21 | 0.48±0.19 |
| Specificity | 0.38±0.14 | 0.57±0.17 | 0.49±0.28 | 0.65±0.11 | 0.53±0.18 | 0.59±0.09 | 0.71±0.05 |
| **RF** |  | | | | | | |
| AUC | 0.48±0.03 (0.41 – 0.55) | 0.60±.0.03 (0.54 – 0.68) | 0.55±0.09 (0.46 – 0.62) | 0.67±0.06 (0.61 – 0.76) | 0.59±0.05 ( 0.50 – 0.65) | 0.58±0.08 (0.49 – 0.64) | 0.49±0.09 (0.42 – 0.57) |
| PPV | 0.36±0.09 | 0.62±0.09 | 0.48±.0.16 | 0.59±0.08 | 0.54±0.08 | 0.59±0.14 | 0.33±0.21 |
| NPV | 0.57±0.05 |  | 0.65±0.11 | 0.73±0.13 | 0.65±0.12 | 0.63±0.13 | 0.61±0.09 |
| Sensitivity | 0.24±0.10 | 0.35±0.11 | 0.30±0.22 | 0.70±0.10 | 0.46±0.15 | 0.42±0.17 | 0.20±0.13 |
| Specificity | 0.72±0.06 | 0.84±0.07 | 0.80±0.12 | 0.64±0.15 | 0.72±0.08 | 0.75±0.17 | 0.79±0.15 |

Table S4: Regression performance of post-treatment Y-BOCS.

|  | fALFF | ReHo | Functional Connectivity | Clinical data | Clinical+fALFF | Clinical+ReHo | Clinical+Functional Connectivity |
| --- | --- | --- | --- | --- | --- | --- | --- |
| **SVR** |  |  |  |  |  |  |  |
| RMSE | 7.27±1.23 | 6.87±1.04 | 7.71±0.61 | 7.18±0.81 | 7.13±1.16 | 6.57±0.99 | 7.20±0.63 |
| R2 | -0.25±0.19 | -0.12±0.19 | -0.56±0.65 | -0.25±0.21 | -0.21±0.18 | -0.02±0.09 | -0.29±0.26 |
| **RF** |  |  |  |  |  |  |  |
| RMSE | 7.06±1.19 | 6.70±1.23 | 7.05±1.21 | 6.88±1.06 | 7.04±1.19 | 6.72±1.18 | 6.10±0.88 |
| R2 | -0.19±0.29 | -0.06±0.17 | -0.20±0.18 | -0.15±0.34 | -0.19±0.29 | -0.07±0.21 | 0.05±0.07 |

Table S5: ROC-analysis using baseline Y-BOCS for predicting remission on the entire sample (n=159) determining the optimal Y-BOCS cut off point with respective sensitivity, specificity, and balanced accuracy.

| Y-BOCS | Sensitivity | Specificity | Balanced Accuracy |
| --- | --- | --- | --- |
| 11.00 | 1.00 | 0.00 | 0.50 |
| 13.50 | 1.00 | 0.01 | 0.51 |
| 15.50 | 1.00 | 0.03 | 0.51 |
| 16.50 | 0.99 | 0.06 | 0.52 |
| 17.50 | 0.97 | 0.06 | 0.51 |
| 18.50 | 0.96 | 0.07 | 0.52 |
| 19.50 | 0.96 | 0.12 | 0.54 |
| 20.50 | 0.95 | 0.19 | 0.57 |
| 21.50 | 0.92 | 0.24 | 0.58 |
| 22.50 | 0.89 | 0.36 | 0.62 |
| 23.50 | 0.87 | 0.46 | 0.67 |
| 24.50 | 0.71 | 0.58 | 0.64 |
| 25.50 | 0.70 | 0.63 | 0.66 |
| 26.50 | 0.60 | 0.72 | 0.66 |
| 27.50 | 0.57 | 0.75 | 0.66 |
| 28.50 | 0.47 | 0.84 | 0.65 |
| 29.50 | 0.42 | 0.85 | 0.64 |
| 30.50 | 0.26 | 0.90 | 0.58 |
| 31.50 | 0.22 | 0.94 | 0.58 |
| 32.50 | 0.11 | 0.97 | 0.54 |
| 33.50 | 0.10 | 0.99 | 0.54 |
| 34.50 | 0.05 | 1 | 0.53 |
| 35.50 | 0.03 | 1 | 0.52 |
| 37.50 | 0.01 | 1 | 0.51 |
| 40.00 | 0.00 | 1 | 0.50 |
